# Impact of biologics and small molecules for Inflammatory Bowel Disease on COVID-19 Related Hospitalization: A Systematic Review and Meta-analysis

**DOI:** 10.1101/2021.10.24.21265344

**Authors:** Fatema Alrashed, Hajer Alasfour, Mohammad Shehab

**Author notes:** **Corresponding author:** Mohammad Shehab, Division of Gastroenterology, Department of Internal Medicine, Mubarak Alkabeer University Hospital, Kuwait University, Aljabreyah, Kuwait. **FINANCIAL SUPPORT:** None. **AUTHOR CONTRIBUTION** Conceptualization: MS Data curation: FA, HA Formal analysis: FA, MS Investigation: MS, FA Methodology: FA, MS, HA Project administration: MS Resources: FA, MS Software: FA, MS Supervision: MS Validation: MS Writing – original draft: FA, HA Writing – review & editing: MS Approval of final manuscript: all authors.

## Abstract

**Background:** The use of biological therapies and small molecules have been a concern for patients with inflammatory bowel disease (IBD) during COVID-19 pandemic. We aim to assess the association between risk of COVID-19 related hospitalization and these agents.

**Method:** A systematic review and meta-analysis of all published studies from December 2019 to September 2021 was performed to identify studies that reported COVID-19 related hospitalization in IBD patients receiving biological therapies or tofacitinib. The risk ratio (RR) was calculated to compare the relative risk of COVID-19 related hospitalization in patients receiving these medications to those who were not, at the time of the study.

**Results:** 18 studies were included. The relative risk of hospitalization was significantly lower in patients with IBD and COVID-19 who were receiving biologic therapy with RR of 0.47 (95% CI: 0.42-0.52, p < 0.00001). The RR was lower in patients receiving anti-TNFs compared to those who did not [RR= 0.48 (95% CI: 0.41-0.55, p < 0.00001)]. Similar finding was observed in patients taking ustekinumab [RR= 0.55 (95% CI: 0.43-0.72, p < 0.00001)]. Combination therapy of anti-TNF and an immunomodulator did not lower the risk of COVID-19 related hospitalization [RR= 0.98 (95% CI: 0.82-1.18, p =0.84]. The use of vedolizumab [RR= 1.13 (95% CI: 0.75-1.73, p =0.56] and tofacitinib [RR= 0.81 (95% CI: 0.49-1.33, p =0.40] was not associated with lower risk of COVID-19 related hospitalization.

**Conclusion:** Regarding COVID-19 related hospitalization in IBD, anti-TNFs and ustekinumab were associated with favorable outcomes. In addition, vedolizumab and tofacitinib were not associated with COVID-19 related hospitalization.

## Introduction

Severe acute respiratory syndrome coronavirus 2 (SARS-CoV-2) is a global pandemic that had evolved shortly after emerging from Wuhan, China in December 2019.^1,2^ A significant number of patients are at risk of hospitalization as results of disease complications, these include vulnerable patients such as elderly and immunocompromised individuals, and those with active malignancy and cardiopulmonary diseases.^3,4^ SARS-COV2 is known to be transmitted through air droplets and aerosols, also airborne transmission can also be considered as a source of transmission.^2^ The ability of SARS-CoV-2 to affect almost any organ across the body is due to the presence of a receptor called angiotensin converting enzyme 2 (ACE2).^5^ Where it is majorly expressed in the alveolar epithelial type II cells in the lungs, the brush border of gut enterocytes and along with the ciliated cells. The intestinal ACE2 receptor is involved in regulating the expression of antimicrobial peptides and promoting the homeostasis of the gut microbiome.^5^ Gastrointestinal (GI) manifestations are common in patients with Coronavirus disease (COVID-19). Additionally, a recent study showed that GI manifestations of COVID-19 are also common in patients with inflammatory bowel disease (IBD).^6^ However, current data shows patients with IBD were not at higher risk for COVID-19 infection.^7^

Patients with IBD are often requiring long-term maintenance medical therapy. Biologic therapies and janus kinase (JAK) inhibitors are commonly used to maintain remission in patients with IBD.^8^ The effect of these medications on COVID-19 outcome is still not fully understood. The Epidemiology of Coronavirus Under Research Exclusion (SECURE-IBD) database is an international registry that was established at the beginning of the pandemic to report the outcomes of COVID-19 in patients with IBD.^9^ To date, it includes outcomes of COVID-19 infection in more than 6000 patients with IBD from 72 countries worldwide. In addition, multiple studies have been performed to evaluate the safety of IBD medications during COVID-19 pandemic with conflicting data.^7,10,11^ However, there is a lack of up to date data because of the rapidity of emerging data. To our knowledge, there was no previous systematic review that looked at individual biologic therapy and risk of hospitalization due to COVID-19 infection. Moreover, this is the first and largest systematic review to include anti-TNF combination therapy and janus kinase inhibitors.

## Methods

Preferred Reporting Items for Systematic Reviews and Meta-Analyses (PRISMA) statement^12^ was used to conduct this systematic review and meta-analysis were conducted as described in the Cochrane Handbook of Systematic Reviews. MOOSE guidelines were also followed.^13^

### Inclusion and exclusion criteria

We included all relevant studies based on predetermined inclusion and exclusion criteria. In terms of types of studies, we included randomized, controlled trials, observational studies, and editorial were included. Furthermore, Surveillance Epidemiology of Coronavirus Under Research Exclusion (SECURE-IBD) data was included. In terms of population, only adult’s patients (age ≥18 years) with inflammatory bowel disease (IBD) and confirmed SARS-CoV 2 infection were included. As for outcomes, any study that reported hospitalization data in IBD patients infected with SARS-CoV 2 and receiving biologic therapy or small molecules Janus Kinase (JAK) inhibitors at the time of the study were included.

We excluded case series, and case reports and any studies that did not have relevant outcome data. In addition, to avoid duplication, any study that reported data from the SECURE-IBD database was excluded. Finally, we also excluded studies that included pediatric patients only (age <18 y).

### Definitions and Outcome Measures

The primary outcome measure was the risk of COVID-19 related hospitalization in patients taking biologic therapy or small molecule JAK inhibitors. Biological agents included tumor necrosis factor antagonists (anti-TNF) adalimumab and infliximab, vedolizumab, and ustekinumab. Association of COVID-19 related hospitalization and individual biologic agents were explored when available. If data on individual biologic agents was not available, we grouped anti-TNFs, vedolizumab and ustekinumab under biologic agents. Data on concurrent use of anti-TNF agents and an immunomodulator (combination therapy) were also extracted.

### Search Strategy

MEDLINE, Embase, Scopus, and Cochrane Central Register of Controlled Trials databases were searched from December 1st, 2019, to September 1st, 2021, using predetermined search terms (Supplementary file table 1). The only restriction applied was English language. In addition, the SECURE-IBD database was searched for relevant data, as well as clinical trials databases [www.clinicaltrials.gov and International Randomized Standard Clinical Trial (IRSCT) Register].

**Table 1.**
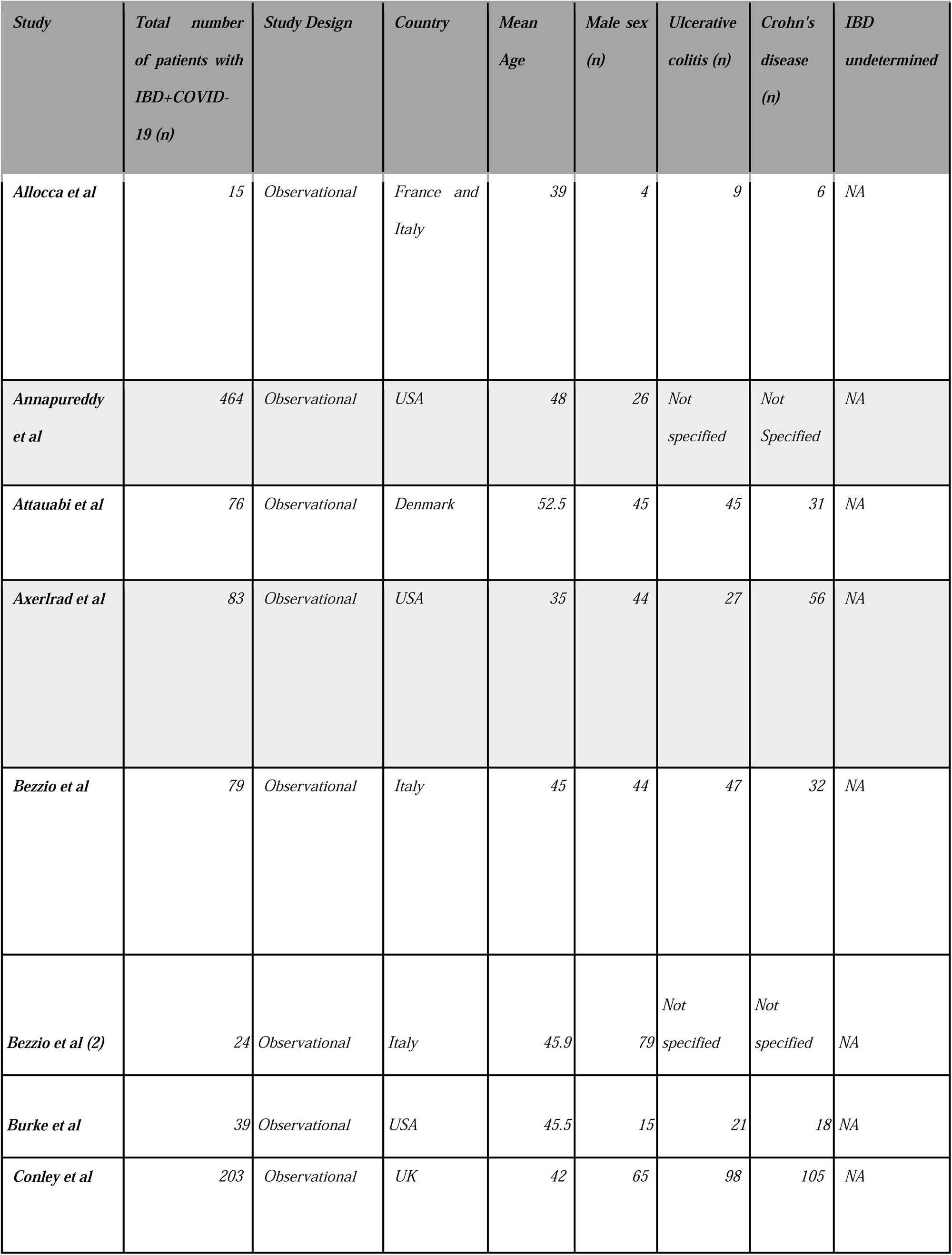
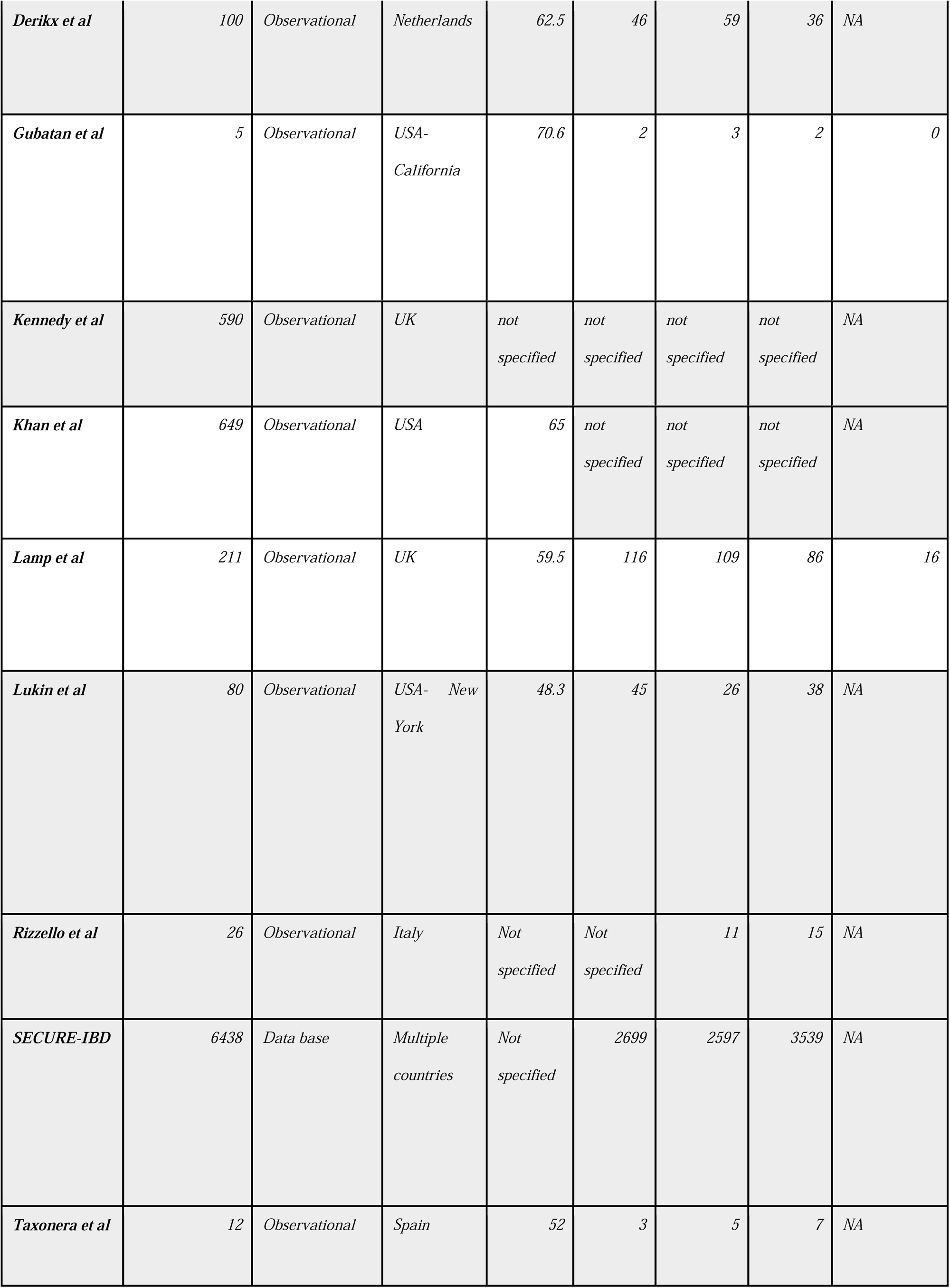
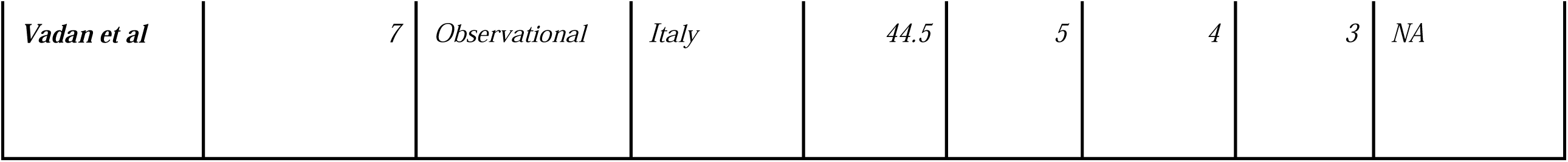
Summary of included studies and patients’ characteristics

English conference proceedings were searched. These include Canadian Digestive Disease Week, Digestive Disease Week, World Congress of Gastroenterology, American College of Gastroenterology, European Crohn’s and Colitis Organization congress, and United European Gastroenterology Week. Furthermore, Google scholar was also searched for unindexed studies. Finally, systematic reviews were also reviewed for relevant studies.

Any relevant titles and abstracts were appraised independently by two authors (FA and HA) and data extraction was performed by the same authors. Differences, if any, were resolved after discussion with a third reviewer (MS).

### Risk of Bias and Study Quality

The modified Newcastle-Ottawa Scale (mNOS)^14^ was used to evaluate the quality of all included studies. This tool allows us to use a scoring system to quantify the quality of studies. A score of 0-3 means the study is judged to be of a low quality, whereas a score of 4-6 means a moderate quality. Finally, any study scoring 7 or 8 is deemed to be of a high quality. This assessment tool uses three domains to appraise the quality of observational studies. These domains are selection, compatibility and outcome.

ROBINS-I for assessing risk of bias in non-randomized studies of interventions^15^ was used for observational studies, whereas, Cochrane risk of bias tool for randomized controlled trials (RoB 2)^16^ was utilized for randomized controlled trials. To evaluate the quality and risk of bias, two authors (MS and FA) worked independently.

### Statistical analysis

The risk ratio (RR) was calculated to compare the relative risk of COVID-19 hospitalization in patients taking biologic therapy and JAK inhibitors to those who were not receiving those medications at the time of the study. Statistical analysis was conducted using Review Manager (RevMan) version 5.3.5 (The Cochrane Collaboration). 95% confidence interval (CI) were estimated using random-effects models given that studies differed in their designs and approach to the research question. The heterogeneity was determined by I^2^ and P value of heterogeneity. In order to quantify the heterogeneity, an I^2^ value of less than 30% indicates low heterogeneity, whereas a moderate heterogeneity was defined as an I^2^ of 30 to 75%. Finally, high heterogeneity was defined as I^2^ value of > 75%.

## Results

### Search Results

From the initial 811 studies identified in the search, 18 studies met criteria for inclusion (Figure 1). This also includes the data extracted from the SECURE-IBD database. All included studies were observational. Six studies were conducted in the United States, five in Italy, three were done in the United Kingdom and the rest were done in multiple countries including France, Spain and Denmark. The main characteristics and findings of the 18 studies are shown in table 1. In total, 9,101 patients with IBD and confirmed COVID-19 diagnosis were included. Mean age was 42 (±10.3) and 3,283 (36%) were male. Among these patients, 3,061 (33.6%) had ulcerative colitis, 3,974 (43.6%) had Crohn’s disease and the remainder did not specify IBD type.

**Figure 1.**
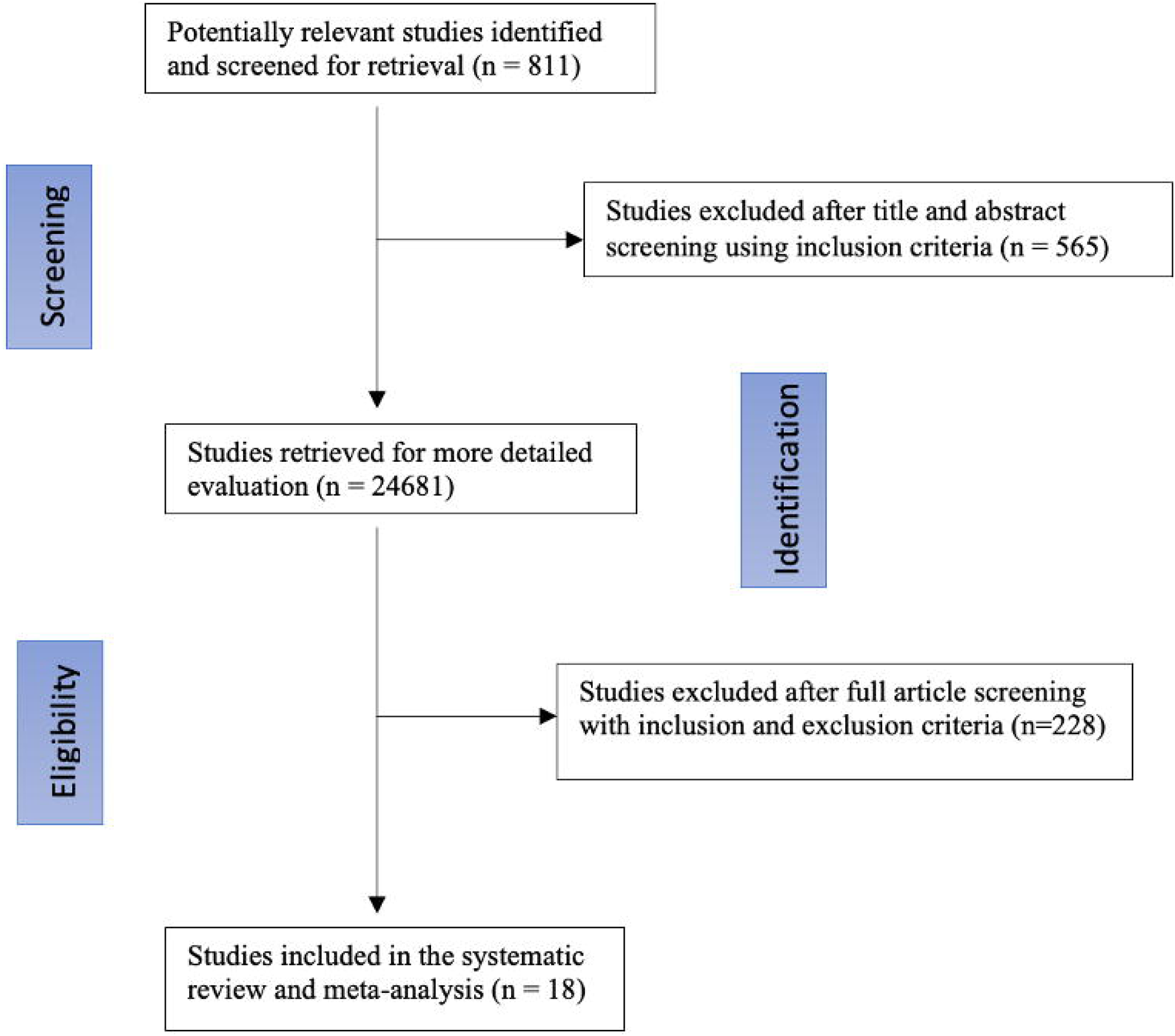
PRISMA diagram showing search strategy

### Outcome

The relative risk of hospitalization was significantly lower in patients with IBD and COVID-19 who were receiving biologic therapy. Specifically, the risk ratio (RR) was 0.47 (95% CI: 0.42-0.52, p < 0.00001). When looking at specific biologic therapy, the relative risk of COVID-19 related hospitalization was lower in patients receiving anti-TNFs compared to those who did not [RR= 0.48 (95% CI: 0.41-0.55, p < 0.00001)]. Similar finding was observed in patients taking ustekinumab [RR= 0.55 (95% CI: 0.43-0.72, p < 0.00001)]. On the other hand, combination therapy of anti-TNF and an immunomodulator did not lower the risk of COVID-19 related hospitalization [RR= 0.98 (95% CI: 0.82-1.18, p =0.84]. Similarly, the use of vedolizumab [RR= 1.13 (95% CI: 0.75-1.73, p =0.56] and tofacitinib [RR= 0.81 (95% CI: 0.49-1.33, p =0.40] was not associated with lower risk of COVID-19 related hospitalization. (Figure 2)

**Figure 2.**
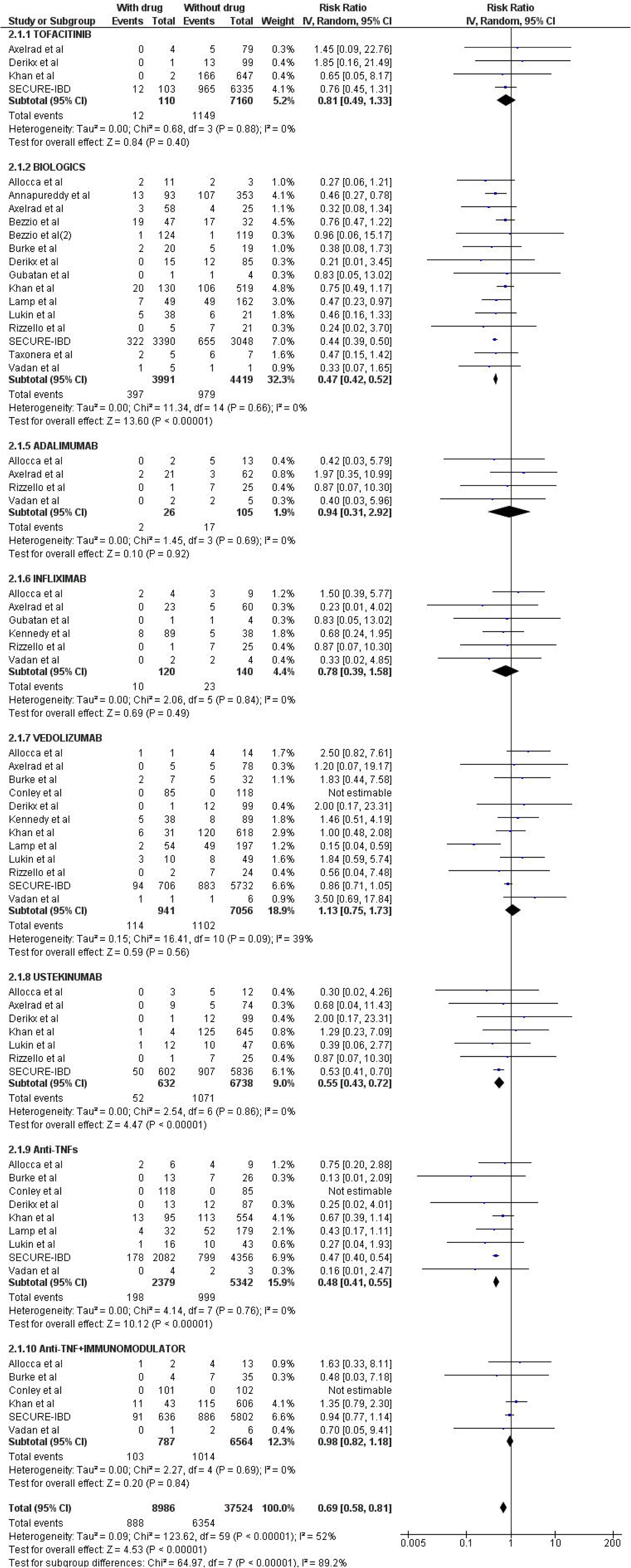
Forest plot showing risk of COVID-19 related hospitalization in patients receiving biological agents and small molecules

### Heterogeneity Assessment, Risk of bias and Quality of Studies

After assessing each study independently, the median mNOS score was 4, with scores ranging from 4 to 6 (see supplemental table 2). In addition, using the Risk of Bias in Non -Randomized Studies - of Interventions (ROBINS-I) tool, most studies were judged to have low risk of bias (see supplemental table 3). Using a random effect model, heterogeneity I^2^ value was low (less than 30%) except for studies evaluating vedolizumab where heterogeneity was medium at 39%.

## Discussion

In this meta-analysis, eighteen studies were identified, evaluating a total of 9,101 patients with inflammatory bowel disease (IBD) and COVID-19. Patients were receiving different biologic therapies including anti-TNFs, vedolizumab, ustekinumab and tofacitinib. Our analysis showed that the risk of COVID-19 related hospitalization significantly decreases in patients taking anti-TNFs, and ustekinumab. Whereas tofacitinib, vedolizumab, and immunomodulators in combination with anti-TNFs were not associated with any negative outcomes.

Our study also showed regarding COVID-19 related hospitalization in IBD, biologics are associated with favorable outcome. Several studies^11,17,18^ described the rationale for trying anti-TNFs therapies in COVID-19. In summary, the mechanism of action of these medications ultimately leads to neutralization of TNF, which plays a major part in the body cytokine response. This response leads to hyperimmune reaction and inflammation in some patients infected with SARS-CoV-2.^19^ Furthermore, the excess inflammatory phase in COVID-19 is characterized by elevated concentrations of serum TNF, interleukin (IL)-6, and IL-8, but relatively little IL-1.^20^ Therefore, it is possible that TNF blockade can reduce COVID-19 hyperinflammation and prevent the need for hospitalization or ICU care. A randomized control trial (CATALYST) is underway in the United Kingdom assessing the use of infliximab in patients hospitalized with severe COVID-19. Additionally, another phase 2 trial conducted in Boston by Tufts Medical Center is also examining the hypothesis that early institution of TNFα inhibitor therapy in patients with severe COVID-19 infections will prevent further clinical deterioration (NCT04425538).

Another notable finding of this meta-analysis is that regarding COVID-19 related hospitalization, ustekinumab, but not vedolizumab, is associated with favorable outcome. Ustekinumab binds to the p40 subunit of interleukin (IL)-12 and IL-23. The binding of this subunit prevents their interaction and attachment with cell surface, preventing IL-12- and IL-23-mediated cell signaling and cytokine production.^21^ Current data propose that interfering with cytokine production will result in dampening of the systemic inflammation caused by SARS-CoV-2 infection that can lead to multiorgan failure and death.^22^ One case report described a case where ustekinumab was not associated with worse outcome in a patient with COVID-19, and attributed this action to the anti-inflammatory effect of the double neutralization of IL-12 and IL-23.^23^ On the other hand, vedolizumab did not exert the same favorable effect, however, it was not associated with increased risk of COVID-19 related hospitalization. Given its gut-specific mechanism of action, vedolizumab does not lead to a significant impact on systemic immunity, therefore, cytokine production may not be significantly reduced to prevent the complications arising from the hyper-immune response caused by SARS-CoV-2 infection described previously. Interestingly, one study compared COVID-19 outcomes among patients on vedolizumab monotherapy to those on anti-TNF monotherapy. Although the difference between the two groups was not significant in terms of severe COVID-19 outcomes, this study found that the risk of hospitalization was 38% more likely to occur with vedolizumab monotherapy compared to anti-TNF monotherapy.^24^ These findings further support the hypothesis that anti-TNF therapies are associated with favorable outcome in COVID-19. Furthermore, one study that enrolled 234 patients^25^ suggested that biologics are not associated with worse COVID outcomes in IBD. The study found that patients taking biologics were 5 times less likely to be diagnosed with the infection, supporting the hypothesis that immunomodulating drugs that dampen cytokine activity have a favorable effect.

Our findings regarding biologic therapy support their continued use and should be reassuring to the large number of IBD patients receiving these agents. Currently, expert are on agreement that active IBD disease leads to more adverse outcomes in patients with SARS-CoV-2 infection compared to medication related immunosuppression, therefore, different professional organizations of gastroenterology are endorsing continuing IBD therapies during the COVID-19 pandemic.^26^ Other recommendations include avoiding immunomodulators if possible, minimizing corticosteroid exposure, and preference for subcutaneous route of drug delivery for initiation of biologic therapy.^27,28^

In this meta-analysis we also found that tofacitinib was not associated with increased risk of COVID-19 related hospitalization. Similarly, a study by Argwal et al found that risk of hospitalization or ICU admission did not differ between tofacitinib-treated patients and other patients.^10^ In addition, a recent randomized controlled trial found that tofacitinib reduces the incidence of death.^29^

This study is the first to comprehensively explore the role of all biological therapies, anti-TNF combination therapy and small molecule inhibitors in COVID-19 related hospitalization. The current study summarizes significantly increased available data overall and for individual biological therapies. A previous systematic review and meta-analysis earlier in the pandemic, included studies up to July 2020. ^30^ also found that biological agents have favorable outcomes in severe COVID-19 disease. However, all biologic therapies were grouped together hence it was difficult to draw a conclusion on the role of different biological agents individually. With a larger sample size, our study found that ustekinumab, but not vedolizumab, is associated with favorable outcome in COVID-19 related hospitalization. Furthermore, this larger sample of IBD patients infected with SARS CoV-2 give more precise estimation of the risk of COVID-19 related hospitalization. In addition, because of the specific and strict outcome measure we have chosen, the heterogeneity was low among all studies. Finally, this study was performed by adhering to the highest of standards including MOOSE guideline and PRISMA statement. Inclusion of good quality studies with detailed extraction of data, and rigorous evaluation of study quality lend great credibility and strength to our systematic review and meta-analysis

Our study limitations include the observational nature of included studies with risk of confounding and selection bias. Furthermore, patient level data were lacking, and insufficient data is available to stratify patients by different factors, including age, disease activity, and socio-economic assessments of the patients. Additional research is needed to ascertain which risk factors play significant roles in causing COVID-19 related hospitalization. Finally, the majority of patients included in our study were extracted from the SECURE-IBD database. One disadvantage of this database is that it may be subject to reporting bias, which means that physicians tend to document severe cases, while the milder cases may remain underreported. Additional research is needed to further evaluate causality between the use of biologic therapies and COVID-19 outcomes.

In conclusion, Regarding COVID-19 related hospitalization in IBD, Anti-TNFs and ustekinumab were associated with favorable outcomes. In addition, vedolizumab and tofacitinib were not associated with COVID-19 related hospitalization.

## Data Availability

All data produced in the present work are contained in the manuscript

## ACKNOWLEDGMENTS

none

