## supplementary documents for "Impact of biologics and small molecules for Inflammatory Bowel Disease on COVID-19 Related Hospitalization: A Systematic Review and Meta-analysis"

**Supplementary Appendix 1: Search strategies**

Supplemental Table 1 | Search strategy

Medline search strategy

(Inflammatory Bowel Disease[Mesh] or Crohn disease[tw] or Crohn Disease[Mesh] or Crohn disease[tw] or Crohn's Enteritis*[tw] or Ulcerative colitis*[tw] or Crohn's Disease[tw] or Crohns Disease[tw] or Ileocolitis*[tw] or Granulomatous Colitis*[tw] or Terminal Ileitis[tw])

AND

(Coronavirus[Mesh] or Coronavirus[tw] or COVID-19[Mesh] or COVID-19[tw] or SARS-COV-2[Mesh] or SARS-COV-2[tw] or severe[tw] or [hospitalization[tw])

AND

("Biologics"[Mesh] or Biologic*[tw] or Biopharmaceutical*[tw] or Biological Drug*[tw] or Biological*[tw] or Biologic Medicine*[tw] or Biologic Pharmaceutical*[tw] or Biologic Drug*[tw] or Biological Medicine[tw] or “Monoclonal Antibodies”[Mesh] or Monoclonal Antibody*[tw] or anti-TNF[tw] or “Tumor Necrosis Factor alpha"[Mesh] or Tumor Necrosis Factor alpha[tw])

Embase search strategy

(Crohn disease.tw. or Crohn's Enteritis.tw. or Regional Enteritis.tw. or Inflammatory Bowel Disease.tw. or Ileocolitis.tw. or Granulomatous Colitis.tw. or Terminal Ileitis.tw. or Regional Ileitides.tw. or Regional Ileitis.tw.)

AND

(Coronavirus.tw. or COVID-19.tw. or SARS-COV-2.tw. OR severe.tw. OR hospitalization.tw. OR mortality.tw.)

AND

(Biologic*.tw. or Biopharmaceutical*.tw. or Biological Drug*.tw. or Biological*.tw. or Biologic Medicine*.tw. or Biologic Pharmaceutical*.tw. or Biologic Drug*.tw. or Biological Medicine.tw. or Monoclonal Antibody*.tw. or adalimumab.tw. or certolizumab.tw. or infliximab.tw. or ustekinumab.tw or vedolizumab.tw. or natalizumab.tw. or Risankizumab.tw)

**Table 2: Quality Assessment**

|  | Selection | | | Comparability | Outcome | | |  |
| --- | --- | --- | --- | --- | --- | --- | --- | --- |
| Study | **Representativeness of sample (maximum: one star)** | **Sample size (maximum: one star)** | **Assessment of the exposure (maximum: one star)** | **Comparability of cohorts on the basis of the design or analysis (maximum: 2 stars)** | **Assessment of the outcome (maximum: one star)** | **Was follow up long enough? (maximum: one star)** | **Adequacy of follow up cohorts (maximum: one star)** | **Total score (maximum: 8 stars)** |
| Allocca et al | * |  | * | * | * |  |  | **** (4) |
| Annapureddy et al | * | * | * | ** | * |  |  | ****** (6) |
| Attauabi et al | * | * |  | ** | * |  |  | ***** (5) |
| Axerlrad et al |  | * | * | * | * |  |  | **** (4) |
| Bezzio et al | * | * |  | ** | * |  |  | ***** (5) |
| Bezzio et al (2) | * | * |  |  | * | * |  | **** (4) |
| Burke et al | * | * |  | * | * |  |  | **** (4) |
| Conley et al | * | * | * |  | * |  |  | **** (4) |
| Derikx et al | * | * |  |  | * |  |  | **** (4) |
| Gubatan et al | * | * | * | ** | * |  |  | ****** (6) |
| Kennedy et al | * | * |  | * | * |  |  | **** (4) |
| Khan et al | * | * | * | * |  | * |  | *****(5) |
| Lamp et al | * | * | * | * |  |  |  | **** (4) |
| Lukin et al | * |  | * | * | * |  | * | ***** (5) |
| Rizzello et al | * | * | * | ** | * |  |  | ******(6) |
| Taxonera et al | * | * |  | ** | * |  |  | ***** (5) |
| Vadan et al | * | * | * |  | * |  |  | **** (4) |

**Table 3: Risk of Bias**

| Study | Risk of bias |
| --- | --- |
| Allocca et al | Low |
| Annapureddy et al | Low |
| Attauabi et al | Low |
| Axerlrad et al | Low |
| Bezzio et al | Low |
| Bezzio et al (2) | Low |
| Burke et al | Low |
| Conley et al | Low |
| Derikx et al | Moderate |
| Gubatan et al | Low |
| Kennedy et al | Low |
| Khan et al | Low |
| Lamp et al | Low |
| Lukin et al | Low |
| Rizzello et al | Low |
| SECURE-IBD | Moderate |
| Taxonera et al | Low |
| Vadan et al | Low |
